# COVID-19 outbreak in Mauritius: Logistic growth and SEIR modelling with quarantine and an effective reproduction number

**DOI:** 10.1101/2020.09.22.20199364

**Authors:** Antoine Géhin, Smita Goorah, Khemanand Moheeput, Satish Ramchurn

**Affiliations:** Lycée des Mascareignes, Helvétia, Saint-Pierre, Mauritius; Department of Medicine, Faculty of Medicine and Health Sciences, University of Mauritius, Reduit, Mauritius; Department of Science, School of Science and Mathematics, Mauritius Institute of Education, Reduit, Mauritius; Department of Physics, Faculty of Science, University of Mauritius, Reduit, Mauritius

## Abstract

**Background and Objectives:** The island of Mauritius experienced a COVID-19 outbreak from mid-March to end April 2020. The first three cases were reported on March 18 (Day 1) and the last locally transmitted case occurred on April 26 (Day 40). An island confinement was imposed on March 20 followed by a sanitary curfew on March 23. Supermarkets were closed as from March 25 (Day 8). There were a total of 332 cases including 10 deaths from Day 1 to Day 41. Control of the outbreak depended heavily on contact tracing, testing, quarantine measures and the adoption of personal protective measures (PPMs) such as social distancing, the wearing of face masks and personal hygiene by Mauritius inhabitants. Our objectives were to model and understand the evolution of the Mauritius outbreak using mathematical analysis, a logistic growth model and an SEIR compartmental model with quarantine and a reverse sigmoid effective reproduction number and to relate the results to the public health control measures in Mauritius.

**Methods:** The daily reported cumulative number of cases in Mauritius were retrieved from the Worldometer website at https://www.worldometers.info/coronavirus/country/mauritius/. A susceptible-exposed-infectious-quarantined-removed (SEIQR) model was introduced and analytically integrated under the assumption that the daily incidence of infectious cases evolved as the derivative of the logistic growth function. The cumulative incidence data was fitted using a logistic growth model. The SEIQR model was integrated computationally with an effective reproduction number (*R*_*e*_) varying in time according to a three-parameter reverse sigmoid model. Results were compared with the retrieved data and the parameters were optimised using the normalised root mean square error (NRMSE) as a comparative statistic.

**Findings:** A closed-form analytical solution was obtained for the time-dependence of the cumulative number of cases. For a small final outbreak size, the solution tends to a logistic growth. The cumulative number of cases was well described by the logistic growth model (NRMSE = 0.0276, *R*^2^ = 0.9945) and by the SEIQR model (NRMSE = 0.0270, *R*^2^ = 0.9952) with the optimal parameter values. The value of *R*_*e*_ on the day of the reopening of supermarkets (Day 16) was found to be approximately 1.6.

**Interpretation:** A mathematical basis has been obtained for using the logistic growth model to describe the time evolution of outbreaks with a small final outbreak size. The evolution of the outbreak in Mauritius was consistent with one modulated by a time-varying effective reproduction number resulting from the epidemic control measures implemented by Mauritius authorities and the PPMs adopted by Mauritius inhabitants. The value of *R*_*e*_ ≈ 1.6 on the reopening of supermarkets on Day 16 was sufficient for the outbreak to grow to large-scale proportions in case the Mauritius population did not comply with the PPMs. However, the number of cases remained contained to a small number which is indicative of a significant contribution of the PPMs in the public health response to the COVID-19 outbreak in Mauritius.

## 1. INTRODUCTION

Infectious diseases have seriously undermined global health for centuries [1, 2]. The rapidity of the spread of the diseases has increased many-fold with the globalisation of air travel [3] and the formation of air traffic hubs [4]. Contextually, the emergence of the zoonotic SARS-CoV-2 virus in Wuhan, Hubei province, China, in late December 2019, [5] and its global spread as a pandemic [6] and manifestation in humans as the highly contagious respiratory disease COVID-19 [7] has been accompanied by large-scale efforts on several fronts to prevent its entry in many territories, contain its spread and mitigate its impact. Entry prevention has, to a large extent, depended on the country-closure of international borders and the quarantining of returning residents. Within countries, spread-containment has been based on active testing and contact tracing and the quarantining of positively-tested persons, lockdowns and the adoption of personal protective measures (PPMs) [8, 9]. Some countries have, however, relied on measures to flatten the epidemic curve, while protecting the more vulnerable persons, so as to alleviate the pressure on their health systems and with the possibility of reaching herd-immunity levels [10].

Mathematical models have significantly contributed to the understanding of the dynamics of infectious diseases. The susceptible-infected-removed (SIR) compartmental model [11] provides a sound epidemiological basis for explaining the S-shape [12] of epidemic curves and explains the concept of herd-immunity from which vaccination-threshold levels can be calculated [11]. The inclusion of an exposed compartment generates the SEIR model which allows the inclusion of incubation and latent periods. More sophisticated models such as agent-based models can work at the level of individuals [13]. Logistic growth models are simpler but have nevertheless been widely used to describe epidemics [14, 15]. All these models have helped in informing and shaping policy decisions on effective responses to the SARS-CoV-2 virus.

Currently, more than 31 million cases of COVID-19 have been recorded worldwide with about 960,000 deaths. In Mauritius, the first three cases were reported on March 18 (Day 1) and the last locally transmitted case occurred on April 26 (Day 40). An island confinement was imposed on March 20 followed by a sanitary curfew on March 23. Supermarkets were closed as from March 25 (Day 8) and reopened on Day 16. There were a total of 332 cases, including 10 deaths, from Day 1 to Day 41. There were no cases for the next 18 days, following which there have been 34 new cases all of whom have been observed from passengers to Mauritius placed in quarantine. Control of the outbreak depended heavily on contact tracing, quarantine measures and the adoption of PPMs such as social distancing, the wearing of face masks and personal hygiene by Mauritius inhabitants. Our objectives were to model and understand the evolution of the Mauritius outbreak using mathematical analysis, a logistic growth model and an SEIR compartmental model with quarantine and a reverse sigmoid effective reproduction number and to relate the results to the public health control measures in Mauritius.

## 2. METHODS

### 2.1. SEIQR model

Compartmental models are widely used in epidemiological analysis [11]. Simple dynamical equations are used to describe the evolution of epidemics. The basic reproduction number and the concept of herd-immunity are easily understood within the framework of, for example, the SIR model where the population is compartmentalised in susceptible, infected and removed classes. Various extensions of the SIR model exist. We have used a susceptible-exposed-infectious-quarantined-removed (SEIQR) compartmental model to model the COVID-19 dynamics with the removed compartment consisting of both the recovered patients and the dead people. A schema of the model together with the dynamical equations used are shown in Figure 1.

**Fig. 1.**
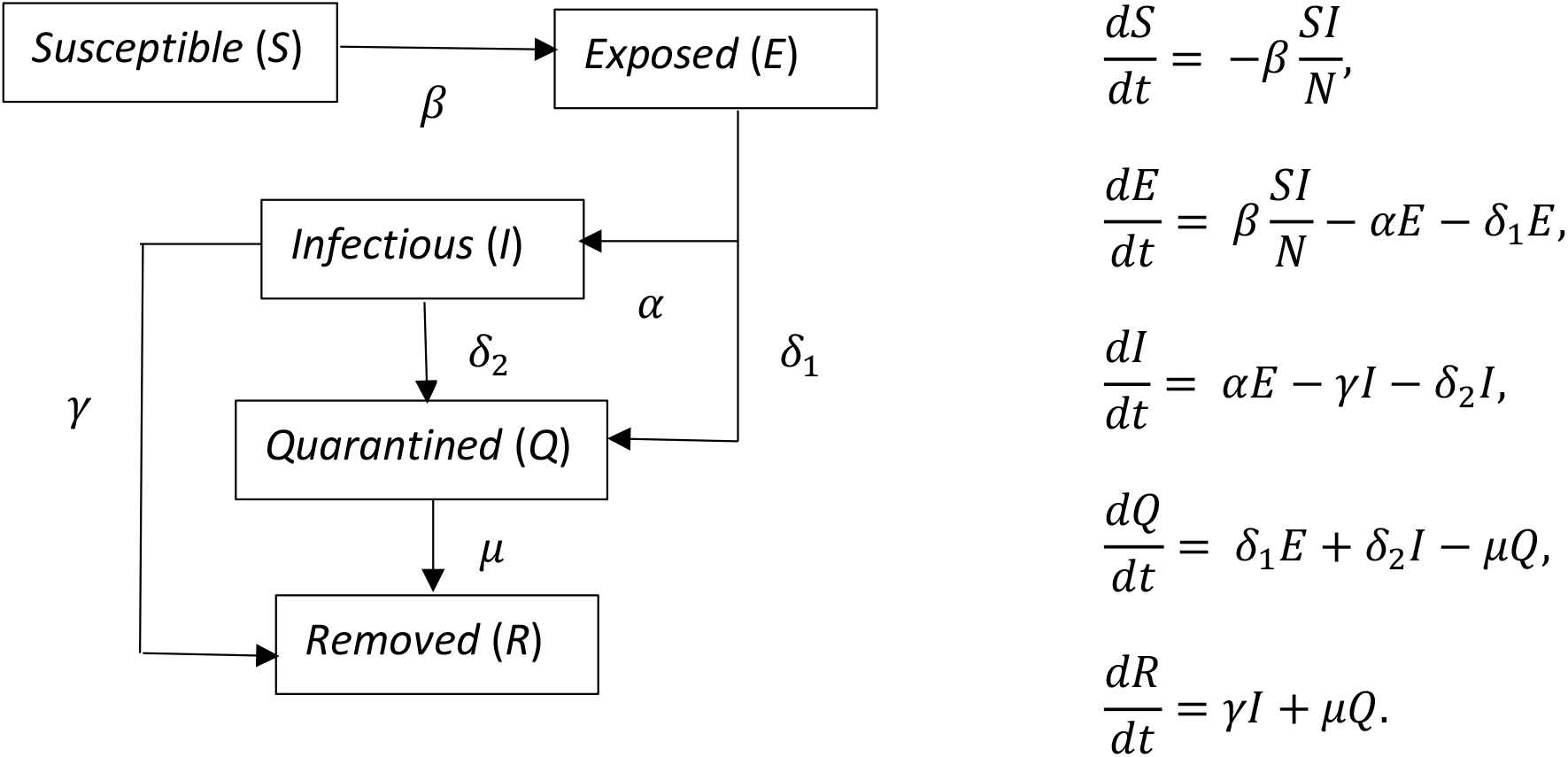
Schema of the SEIQR model and the dynamical equations used.

The time variable, infection rate, removal rates from the infectious and the quarantine compartments are represented by *t, β, γ* and *μ* respectively and 1/*α* represents the average latent period. The removal rates from the exposed and infectious compartments due to quarantine are represented by *δ*_1_and *δ*_2_ respectively. It was also assumed that *S* + *E* + *I* + *Q* + *R* = *N*, where the total population *N* is constant.

### 2.2. Mathematical analysis

The dynamical equation for the cumulative number of cases *N*_*c*_(*t*) = *E* + *I* + *Q* + *R*,

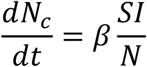

where *N* is the total population, was analytically integrated with the substitution *S* = *N* − *N*_*c*_ and the solution considered under the assumption that the incidence of infectious cases, *βI*, evolved as the derivative of the logistic growth function:

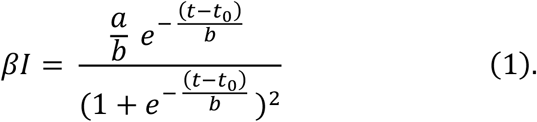

### 2.3. Logistic growth modelling

The logistic growth model was introduced by Verhulst in 1836 [16] as a model of population growth in a limited environment where if the population at time *t* is *N*(*t*) then

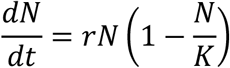

is the logistic equation where *r* is the growth rate, *K* is the carrying capacity of the environment and (1 − *N*/*K*) is a self-regulatory term which limits the growth of the population. The solution of the logistic equation is the logistic growth function

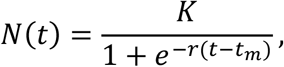

where

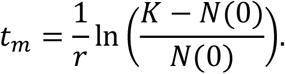

The growth curve is S-shaped if *N*(0) < *K*/2. Several extensions of the Verhulst model have been proposed to address deviations from the symmetric shape of the logistic curve [15]. Epidemiologically, the logistic equation is an exact solution of the susceptible-infected-susceptible (SIS) compartmental model for the infected cases where it is assumed that individuals who recover become immediately susceptible again because the disease confers no immunity against reinfection [17].

We have modelled the cumulative number of COVID-19 cases, *N*_*c*_(*t*), in Mauritius using the logistic growth function

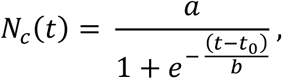

where *a* is the maximum cumulative case incidence (final outbreak size) and 1/*b* is the intrinsic growth rate during the exponential phase, *t*_0_ is the inflection point, the time where maximum number of cases per day occur. Computations were done using Python (with SciPy library) and MATLAB.

### 2.4. SEIQR modelling

The SEIQR dynamical equations were integrated using MATLAB. It was assumed that the effective reproduction number (*R*_*e*_) varied in time according to the reverse sigmoid model

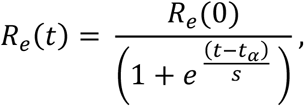

with the inflection point and scale parameter now represented by *t*_*α*_ and *s* respectively. Referring to Figure 1, the infection rate was *β*(*t*) = *R*_*e*_/(*γ* + *δ*_2_). As initial conditions, we used *S*(0) = *N* and 3 persons in the infectious compartment and none in the others. The values of *α, δ*_1_, *γ, δ*_2_, *μ* and *N* were respectively 0.25 day^−1^, 0.1 day^−1^, 0, 0.5 day^−1^, 1/14 day^−1^ and 1.22 *×* 10^6^. The value of *γ* was taken as zero because it was assumed all infectious persons had been diagnosed as such and quarantined before moving to the removed compartment.

Simulations were performed for 2.5 ≤ *R*_*e*_(0) ≤ 6.0 over a range of *t*_*α*_ and *s* values. For every *R*_*e*_(0), *t*_*α*_ and *s*, the computed cumulative number of cases were compared with the reported cumulative incidence data using the normalised root mean square error (NRMSE) as a comparative statistic. The NRMSE was defined as

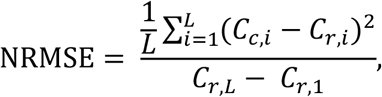

where *C*_*c,i*_ and *C*_*r,i*_ are respectively the cumulative number of computed and reported cases on the *i*^*th*^ day. The last day is denoted by *L*. This process was repeated for all the values of *R*_*e*_(0) so as to obtain the optimal values of *R*_*e*_(0), *t*_*α*_ and *s* which gave the minimum NRMSE with the additional constraint that *R*_*e*_ should not decrease by more than 10% on the first day of the simulation so as to include the realism of the limits of the efficacy of control measures.

## 3. RESULTS

### 3.1. Mathematical analysis

From the dynamical equations,

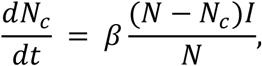

where *N* is the total population, *N*_*c*_ = *E* + *I* + *Q* + *R* and *S* = *N* − *N*_*c*_.

On integration and assuming that *βI* evolves as the derivative of the logistic growth function as in Equation 1, it follows that for *a ≪ N* and *t*_0_ significantly larger than *b*,

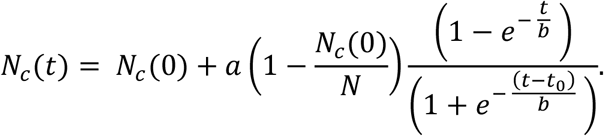

For small *t* and *N*_*c*_(0) ≪ *N*,

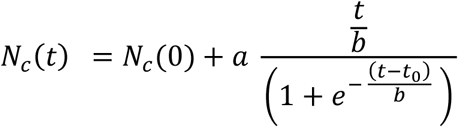

which corresponds to a linear growth modulated by a logistic function, whereas when *t* becomes large compared to *b*

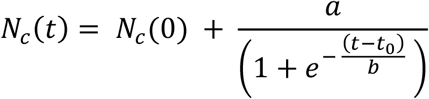

which corresponds to a logistic growth. Details are given in the Appendix.

### 3.2. Logistic growth modelling

Corresponding to the basic premise of the logistic model, as of March 20, the cumulative COVID-19 cases curve of Mauritius can be described by a sigmoid curve with a single turning point (Figure 2). High correlations between the observed and predicted incidence were found (NRMSE = 0.0276, *R*^2^ = 0.9945).

**Fig. 2.**
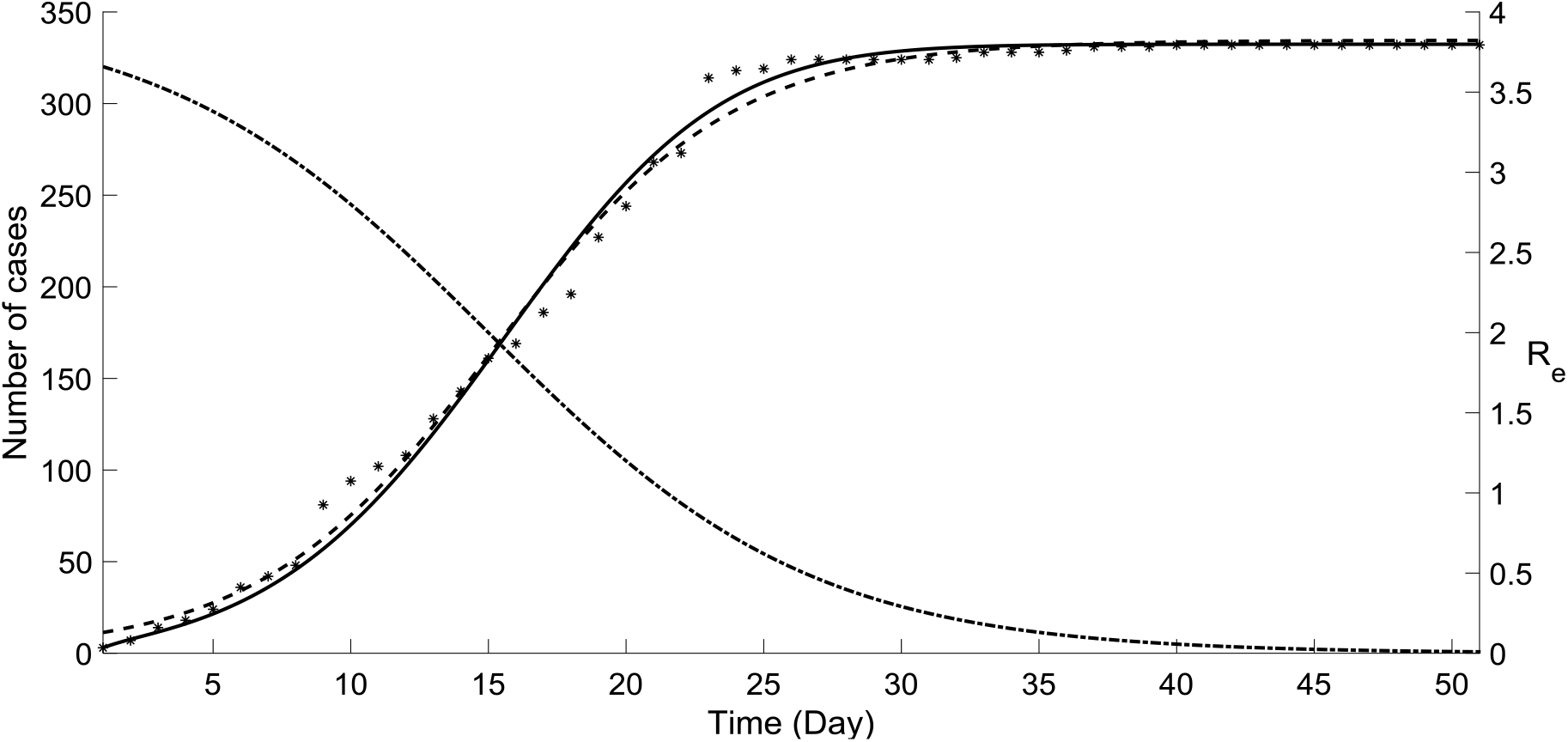
Reported cumulative number of cases in Mauritius (*) and graphical representation of the modelling results: logistic growth (---), SEIQR simulation (full curve) using the optimal effective reproduction number (– · – · –) with the parameter values given in the text.

### 3.3. SEIQR modelling

The optimal values of *R*_*e*_(0), *t*_*α*_ and *s* which gave the minimum NRMSE under the 10% constraint were found to be 4.0, 14.0 and 5.9 respectively. The variation of the reciprocal of the NRMSE for *R*_*e*_(0) = 4.0 is shown in Figure 3 as an intensity colormap. The intensity peak corresponds to the values of *t*_*α*_ and *s* for which the NRMSE is a minimum.

**Fig. 3.**
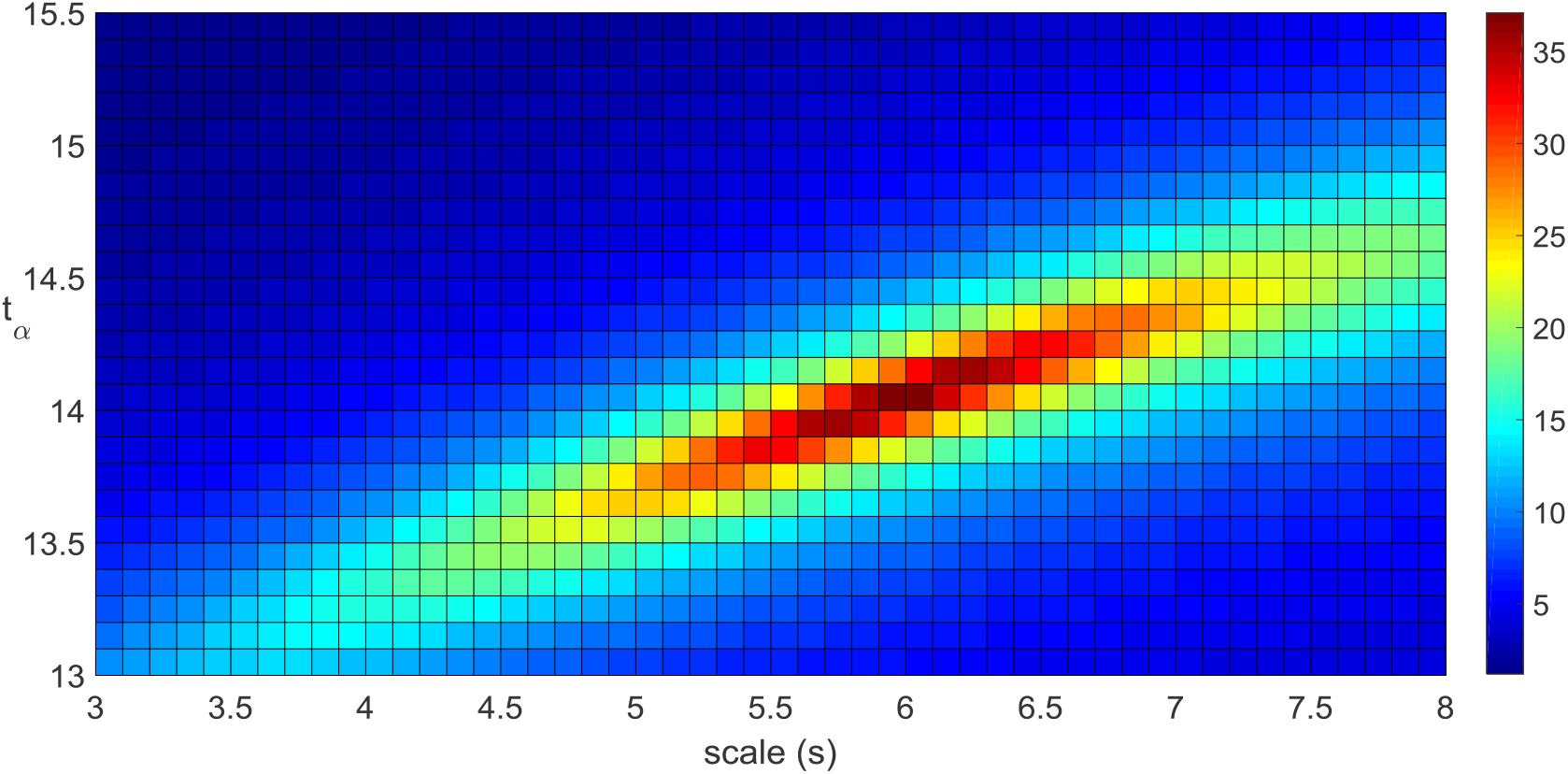
Comparison of the computed cumulative number of cases using the SEIQR model with the reported number of cases for *R*_*e*_(0) = 4.0 and varying *t*_*α*_ and *s*. The colour plot shows the reciprocal of the NRMSE which was used as the comparative statistic.

The computed best-fit cumulative incidence curve (NRMSE = 0.0270, *R*^2^ = 0.9952) obtained using the optimal parameter values in the SEIQR simulations is shown in Figure 2. The optimal *R*_*e*_(*t*) is also shown. The value of *R*_*e*_ on the day of the reopening of supermarkets (Day 16) was found to be approximately 1.6.

## 4. DISCUSSION

The prediction of final epidemic sizes remains a challenging problem. Logistic models may require the presence of the inflection point in the reported data for good estimates. They, however, do not come with the epidemiological principles anchored in compartmental models such as the SEIR model, where, for example, the inflection of the epidemic curve can be explained using herd-immunity concepts. For the COVID-19 outbreak in Mauritius with a small final outbreak size, though, the inflection arises from a time-varying decreasing effective reproduction number resulting from control measures and compliance to PPMs and are not due to herd-immunity effects. SEIR models, such as the one used here, are themselves limited by the homogeneous mixing assumption and by the accuracy of the parameters used. Also, asymptomatic cases were not included. Additionally, the errors in the NRMSE calculations may not be normally distributed with the consequence that the mean may not be a complete statistic. However, in spite of these limitations, the models provide a good description of and insight in the evolution of the Mauritius outbreak.

In particular, we have shown analytically how the solution of a simple SEIQR model tends to a logistic equation under the assumption that the final outbreak size is small, that the number of cases at the point of inflection is sufficiently large and that the incidence of infectious cases evolves as the derivative of the logistic growth function. Computationally, a good fit was obtained for the epidemic curve of the reported cumulative number of cases using a three-parameter logistic growth model. An equally good fit was also computationally obtained using the SEIQR model with an optimal reverse sigmoid time-varying effective reproduction number. Importantly, the value of *R*_*e*_ ≈ 1.6 on the day of the reopening of supermarkets was sufficient for the outbreak to grow to large-scale proportions in case of the non-compliance of the population to the PPMs. However, the number of cases remained contained to a small number which is indicative of a significant contribution of the PPMs in the public health response to the COVID-19 outbreak in Mauritius.

Mauritius has successfully contained the outbreak of COVID-19 on its territory. Its response strategy to the SARS-CoV-2 virus has been centered on early virus entry prevention, controlling and containing the spread of the virus, principally through an intensive contact tracing, testing, strict quarantine measures and enlisting public cooperation in the public health response so that PPMs are strictly adhered to, and on impact mitigation measures. The response has benefited from the lessons learnt in dealing with recent chikungunya [18] and dengue fever outbreaks [19] particularly with respect to the need for control measures at the frontiers, rapid contact tracing and intervention and an efficient communication strategy. There have, however, been 10 deaths and these have been partly attributed to the high level of comorbidities such as diabetes and cardiovascular diseases in the Mauritius population which are known to be strong risk factors for severe complications with COVID-19 [20, 21]. As the island reopens its international borders to all incoming passengers, but with a strict compliance to a two-week quarantine period as from arrival accompanied by PCR tests, periodic recalls for strict compliance to the PPMs are of the essence, while surveillance and contact tracing capabilities need to be reinforced and upscaled, a process which could be underpinned by global collaborations within a One World One Health approach.

## Data Availability

The daily reported cumulative number of cases in Mauritius were retrieved from the Worldometer website at https://www.worldometers.info/coronavirus/country/mauritius/

## Contributions

AG did the logistic modelling. SG, KM and SR did the SEIQR modelling. SR did the mathematical analysis. All authors have contributed to, read, and approved the manuscript. Authors appear in alphabetical order.

## APPENDIX

From the dynamical equations,

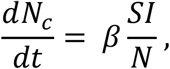

where *N* = (*S* + *E* + *I* + *Q* + *R*) is the total population and *N*_*c*_ = *E* + *I* + *Q* + *R*.

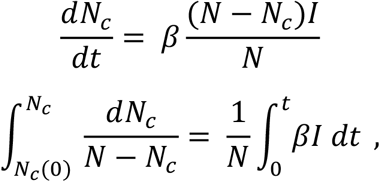

where *β* = *β*(*t*).

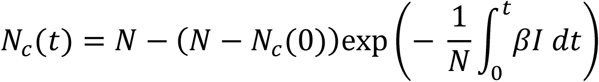

On integration and assuming that *βI* evolved as the derivative of the logistic growth function, i.e.

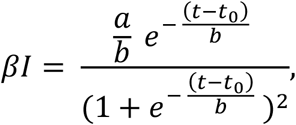

and noting that

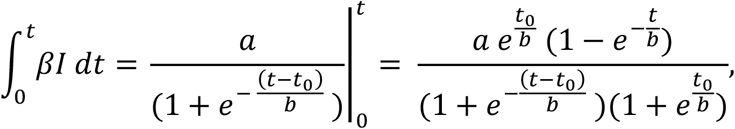

we get the closed-form solution

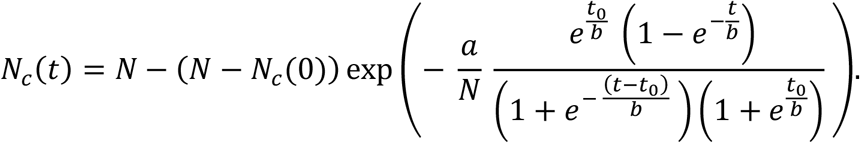

For *a ≪ N* and *t*_0_ significantly larger than *b*,

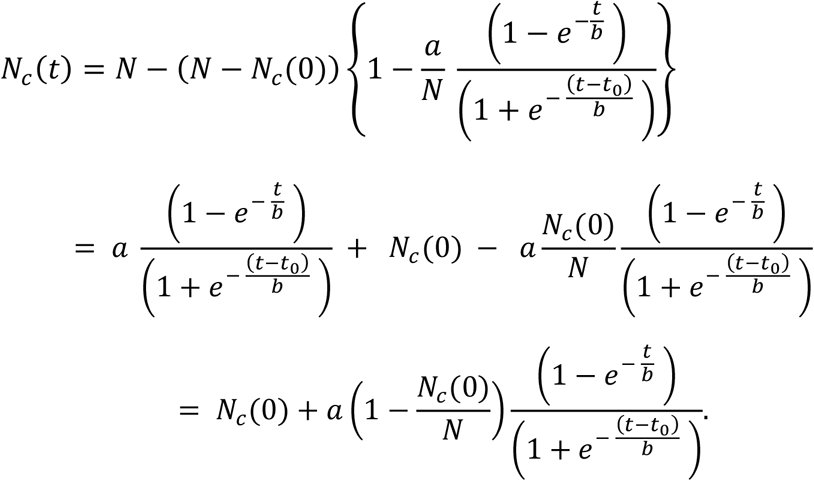

For small *t* and *N*_*c*_(0) ≪ *N*,

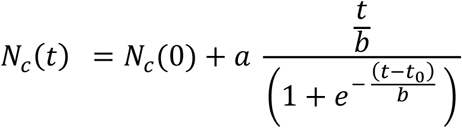

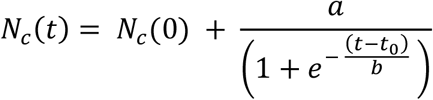

which corresponds to a logistic growth.

